# Characterizing the blood stage antimalarial activity of pyronaridine in healthy volunteers experimentally infected with *Plasmodium falciparum*

**DOI:** 10.1101/2023.09.13.23295466

**Authors:** Bridget E. Barber, Rebecca Webster, Adam J. Potter, Stacey Llewellyn, Nischal Sahai, Indika Leelasena, Susan Mathison, Karsten Kuritz, Julia Flynn, Stephan Chalon, Anne Claire Marrast, Nathalie Gobeau, Joerg J. Moehrle

## Abstract

Although pyronaridine has been used to successfully treat malaria for many years, its antimalarial activity in humans has not been completely characterized. This volunteer infection study aimed to determine the pharmacokinetic/pharmacodynamic (PK/PD) relationship of pyronaridine in healthy malaria naïve adults. Volunteers were inoculated with *Plasmodium falciparum* 3D7-infected erythrocytes on day 0 and different single oral doses of pyronaridine were administered on day 8. Parasitemia, and concentrations of pyronaridine in whole blood were measured and standard safety assessments performed. Curative artemether-lumefantrine therapy was administered if parasite regrowth occurred, or on day 47±2. Outcomes were parasite clearance kinetics, PK and PK/PD parameters from modelling. Ten participants were inoculated and administered 360 mg (n=4), 540 mg (n=4), or 720 mg (n=1) pyronaridine. One participant was withdrawn without receiving pyronaridine. Time to maximum pyronaridine concentration after dosing was 1-2 hours and the elimination half-life was 8-9 days. A parasite clearance half-life of approximately 5 hours was calculated for all dose levels. Parasite regrowth occurred after dosing with 360 mg (4/4 participants) and 540 mg (2/4 participants). Key efficacy parameters of pyronaridine including the minimum inhibitory concentration (MIC: 5.5 ng/mL) and minimum parasiticidal concentration that leads to 90% of maximum effect (MPC_90_: 8 ng/mL) were derived from the final PK/PD model. Adverse events considered related to pyronaridine were predominantly mild to moderate gastrointestinal symptoms. There were no serious adverse events. Data obtained in this study will support the use of pyronaridine in new antimalarial combination therapies by informing partner drug selection and dosing considerations.

## INTRODUCTION

Malaria continues to be responsible for a large global health burden, with an estimated 247 million cases and 619,000 deaths in 2021 [1]. The emergence of artemisinin-resistant *Plasmodium falciparum* in Southeast Asia [2], and more recently in East Africa [3], threatens the utility of the current first line artemisinin combination therapies (ACTs) and progress toward malaria eradication. The development of novel antimalarial therapies is an important strategy to combat the spread of resistance and there are currently significant efforts to progress several promising new antimalarial compounds through clinical development [4]. However, the progress of a new compound from bench to bedside is typically a lengthy process and additional strategies are required to control malaria in the nearer term. Optimizing the use of existing non-artemisinin antimalarial drugs is one such strategy that has potential to reduce malaria morbidity and mortality and slow the spread of antimalarial resistance.

Pyronaridine is an antimalarial compound first developed in China in the 1970’s and has been shown to have potent activity towards blood-stage *P. falciparum*, including isolates resistant to chloroquine, quinine, amodiaquine, pyrimethamine or mefloquine [5]. The antimalarial activity of pyronaridine is thought to occur primarily due to inhibition of hemozoin formation, which results in accumulation of toxic hematin in the parasite food vacuole [6]. Pyronaridine has been used as a single agent for the treatment of malaria in China for over 30 years and was subsequently developed as a fixed-dose combination therapy with artesunate (an artemisinin derivative) for the treatment of acute uncomplicated *P. falciparum* malaria and blood stage *P. vivax* (Pyramax®: pyronaridine-artesunate). *Pyramax* is currently registered in several countries in Africa and Asia, has received positive opinion through article 58 by the European Medicines Agency [7], is on the World Health Organization (WHO) Model Lists of Essential Medicines for adults and children, and is recommended by the current WHO guidelines for malaria [8]. The recommended *Pyramax* dosing regimen for patients ≥65 kg is 720 mg pyronaridine/ 240 mg artesunate daily for three consecutive days.

Despite the fact that pyronaridine has been used to successfully treat malaria for many years, either in monotherapy or in combination with other antimalarial drugs [5], its antimalarial activity in humans has not been completely characterized. Determining the relationship between pyronaridine pharmacokinetics (PK) and pharmacodynamic (PD) antimalarial activity in humans would inform efforts to broaden its use in new combination therapies. Combination therapies are the focus of antimalarial drug development to reduce the risk of selecting for resistant mutants and to target multiple stages of the parasite lifecycle including the transmissible gametocytes [9]. Further, using a combination of two or more drugs is typically considered a prerequisite for achieving cure without the need for an extended dosing regimen, which would pose compliance challenges. In the case of ACTs, the fast-acting artemisinin component rapidly reduces the parasite burden, while a partner drug with a more moderate rate of action but longer elimination half-life is relied upon to clear residual parasites and prevent recrudescence. The rate of action of pyronaridine in clearing malaria parasitaemia in humans is unclear; however clinical pharmacokinetic studies in malaria patients have demonstrated that it has a relatively long elimination half-life of 13.2 days in adults and 9.6 days in children [5] and thus may be able to offer extended protection from recrudescence.

Volunteer infection studies (VIS) using the induced blood stage malaria (IBSM) model have previously successfully characterized the PK/PD relationship of several experimental antimalarial compounds as well as antimalarial drugs currently in use [10–13]. The IBSM model involves intravenous inoculation of healthy malaria naïve adult participants with blood-stage parasites and subsequent administration of the test antimalarial. Frequent blood sampling is performed to measure parasitaemia and drug concentration kinetics over the course of the study, thus providing data for PK/PD modelling analyses. Such VIS have been shown to accurately predict the activity of investigational antimalarials administered as monotherapy in studies in endemic populations [14, 15].

The primary objective of the current study was to determine the PK/PD relationship of pyronaridine following administration in healthy adult participants using the *P. falciparum* IBSM model. Data obtained in this study will support the use of pyronaridine in new antimalarial combination therapies by informing partner drug selection and dosing considerations.

## METHODS

### Study design and participants

This was an open label, randomized, clinical trial using the IBSM model. Healthy malaria naïve males and females (non-pregnant, non-lactating) aged 18-55 years were eligible for inclusion (Text S1). The study was conducted at the University of the Sunshine Coast Clinical Trials Unit (Morayfield, Australia) and was registered on ClinicalTrials.gov (NCT05287893). This study was approved by the QIMR Berghofer Human Research Ethics Committee (P3800). All participants gave written informed consent.

### Procedures

Participants were inoculated intravenously with approximately 2800 viable *P. falciparum* 3D7 infected erythrocytes on day 0. Parasitemia was monitored throughout the study by quantitative PCR (qPCR) targeting the gene encoding *P. falciparum* 18S rRNA [16]. A single oral dose of pyronaridine in tablet form (Shin Poong Pharmaceutical Co., Ltd, Seoul, South Korea) was administered on day 8 after an overnight fast. Participants were randomized to a dose group on day 8. The randomization schedule was generated using SAS. No blinding was performed.

Whole blood concentrations of pyronaridine were measured by liquid chromatography tandem mass spectrometry using a validated method. Participants received a curative course of artemether-lumefantrine (Riamet®, Novartis Pharmaceuticals) if asexual parasite regrowth was detected (≥5,000 parasites/mL), or on day 47±2. The presence of female gametocytes, male gametocytes and ring stage parasites in participants’ blood samples was detected at select time points using qRT-PCR targeting *pfs25*, *pfMGET* and *SBP-1* parasite transcripts respectively [17, 18]. The protocol specified that a single 45 mg dose of primaquine (Primacin®, Boucher & Muir Pty Ltd) could be administered after artemether-lumefantrine treatment (cohort 1) or any time after pyronaridine dosing (cohort 2) to clear gametocytemia. Safety assessments, including monitoring of adverse events (AEs), vital signs, hematology and biochemistry, physical examination, and electrocardiographs, were performed throughout the study.

### Outcomes and Statistical analysis

Analysis of parasite clearance kinetics following pyronaridine was performed in R statistical package version 4.1.3 (2022-03-10). The parasite reduction ratio over a 48-hour period (PRR_48_) and parasite clearance half-life (PCt_1/2_) were estimated using the slope of the optimal fit for the log-linear relationship of the parasitemia decay [19]. Non-compartmental pharmacokinetic analysis was performed using Phoenix version 8.3.

Pharmacokinetic (PK) and pharmacokinetic/pharmacodynamic (PK/PD) modelling and simulations were conducted within R (3.6.3) combined to the IQRtools package (1.9.0) and Monolix (MLX2019R1). PK/PD analyses were performed using non-linear mixed effects models. A population PK model was developed to obtain individual PK parameter estimates that adequately described the observed individual PK profiles. Model selection was based on the Bayesian Information Criterion (BIC), precision of parameter estimates, goodness of fit and visual evaluation of the individual blood profiles. The PK/PD model was then built using the individual PK parameter estimates as regression parameters. Parasitaemia data were log transformed and observations below the lower limit of quantification were censored. Additionally, data points where gametocytes were estimated to exceed 10% of the total parasitaemia count were removed from the data set since only the asexual parasites were of interest for the PD model and the count of gametocytes is not precise enough to back-calculate the number of asexual parasites from the number of total parasites. Precision of parameter estimates, visual evaluation of the individual parasitemia profiles and goodness of fit plots ensured that the data were adequately described by the final PK/PD model.

Key efficacy parameters including the concentration that leads to 50% of maximum effect (EC_50_), maximum effect (E_max_), parasite reduction rate over 48 hours (PRR_48_), parasite elimination half-life, minimum inhibitory concentration (MIC), and minimum parasiticidal concentration (concentration that leads to 90% of maximum effect) (MPC_90_) were derived from the final PK/PD model.

## RESULTS

### Participants

The study was conducted from 04 April 2022 to 26 September 2022. Ten participants were enrolled across two cohorts and intravenously inoculated with *P. falciparum* on day 0 (Figure 1). Participants were healthy malaria-naïve males (n=7) or females (n=3) aged 19 to 52 years (Table 1). One participant enrolled in cohort 1 tested positive for COVID-19 on day 6 and therefore was not dosed with pyronaridine. This participant instead received immediate definitive antimalarial treatment with artemether-lumefantrine. The remaining 9 participants were randomized to a pyronaridine dose group on day 8 (360 mg, n=4; 540 mg, n=4; or 720 mg, n=1). Doses were selected to optimally characterize the PK/PD relationship. All 9 randomized participants received the allocated dose of pyronaridine, completed the study and were included in the analysis of study endpoints.

**Figure 1.**
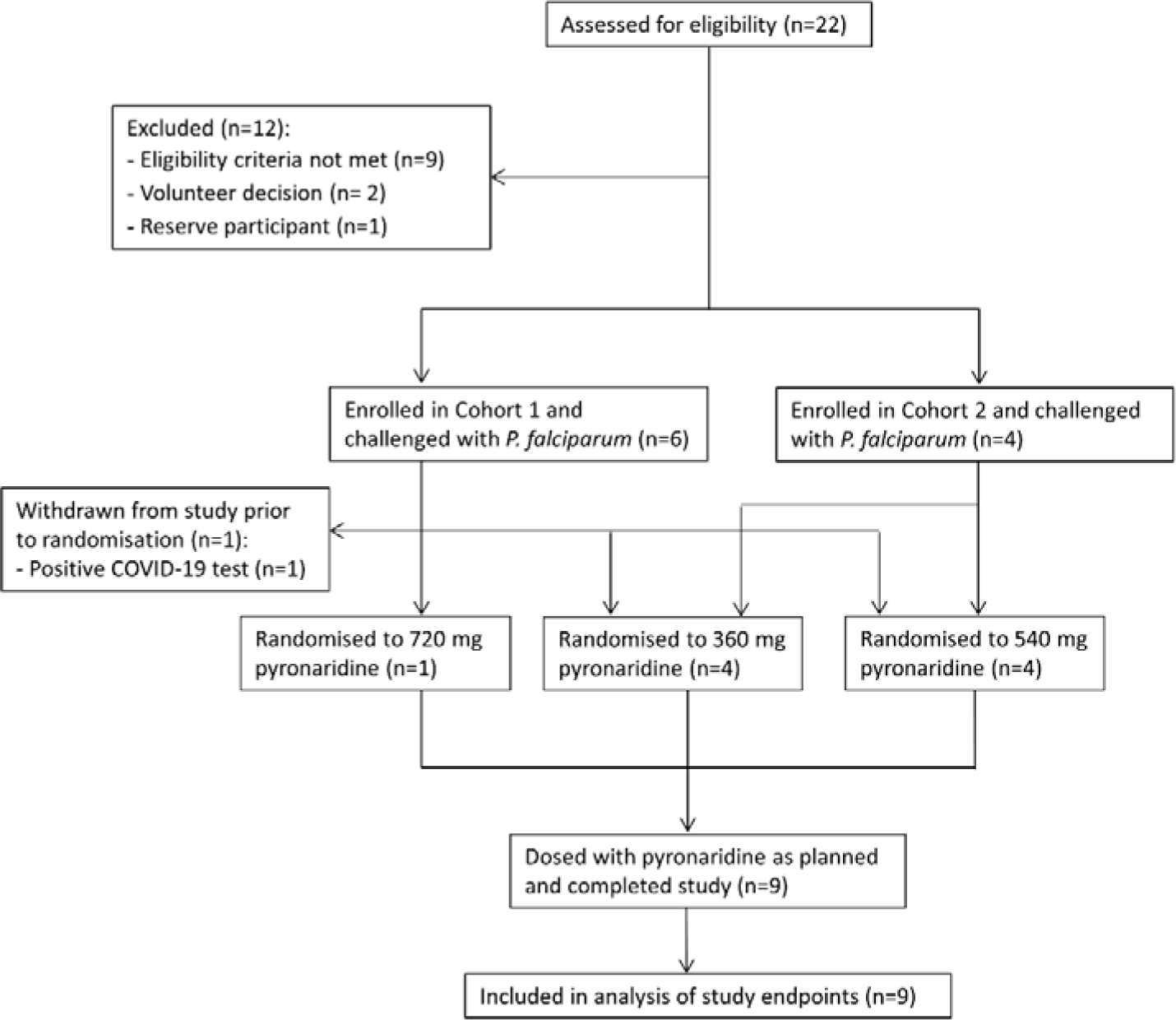
Trial profile. Eligible participants were enrolled in one of two cohorts and challenged with blood stage *P. falciparum*. Participants were randomized within each cohort to a pyronaridine dose group on the day of dosing (8 days post-challenge). One participant was withdrawn from the study after inoculation with malaria but prior to scheduled dosing with pyronaridine. This participant was excluded from analysis of study endpoints.

**Table 1.** Demographic and baseline characteristics of participants.

|  |  | **Pyronaridine 360 mg** | **Pyronaridine 540 mg** | **Pyronaridine 720 mg** | **Total** |
| --- | --- | --- | --- | --- | --- |
|  |  | **(N=4)** | **(N=4)** | **(N=1)** | **(N=10\*)** |
| Sex [n (%)] | Female | 0 (0) | 1 (25) | 1 (100) | 3 (30) |
|  | Male | 4 (100) | 3 (75) | 0 (0) | 7 (70) |
| Race [n (%)] | White | 2 (50) | 4 (100) | 1 (100) | 8 (80) |
|  | Asian | 1 (25) | 0 (0) | 0 (0) | 1 (10) |
|  | Other | 1 (25) | 0 (0) | 0 (0) | 1 (10) |
| Age [years] | Mean (range) | 30.8 (20-37) | 36 (19-52) | 19 (19-19) | 32.8 (19-52) |
| Body weight [kg] | Mean (range) | 77.6 (68.7-81.9) | 79.1 (57.5-104) | 87.9 (87.9-87.9) | 78.3 (57.5-104) |
| Body mass index [kg/m<sup>2</sup>] | Mean (range) | 25.5 (23-29) | 25.8 (19-30) | 25.0 (25-25) | 25.4 (19-30) |

\*Includes one participant who was inoculated with malaria but was withdrawn from the study without being dosed with pyronaridine.

### Parasitemia and drug exposure

The progression of parasitemia following intravenous inoculation was consistent between participants up to day 8, when a single dose of pyronaridine was administered (Figure 2). Dose-related increases in pyronaridine exposure (maximum concentration [C_max_] and area under the concentration-time curve from time 0 to the last measured time point [AUC_0-last_]) were observed across the dose range (Table 3). Absorption was rapid, with time to maximum concentration (t_max_) approximately 1-2 h, while a relatively long elimination half-life (t_1/2_) of approximately 8-9 days was observed.

**Figure 2.**
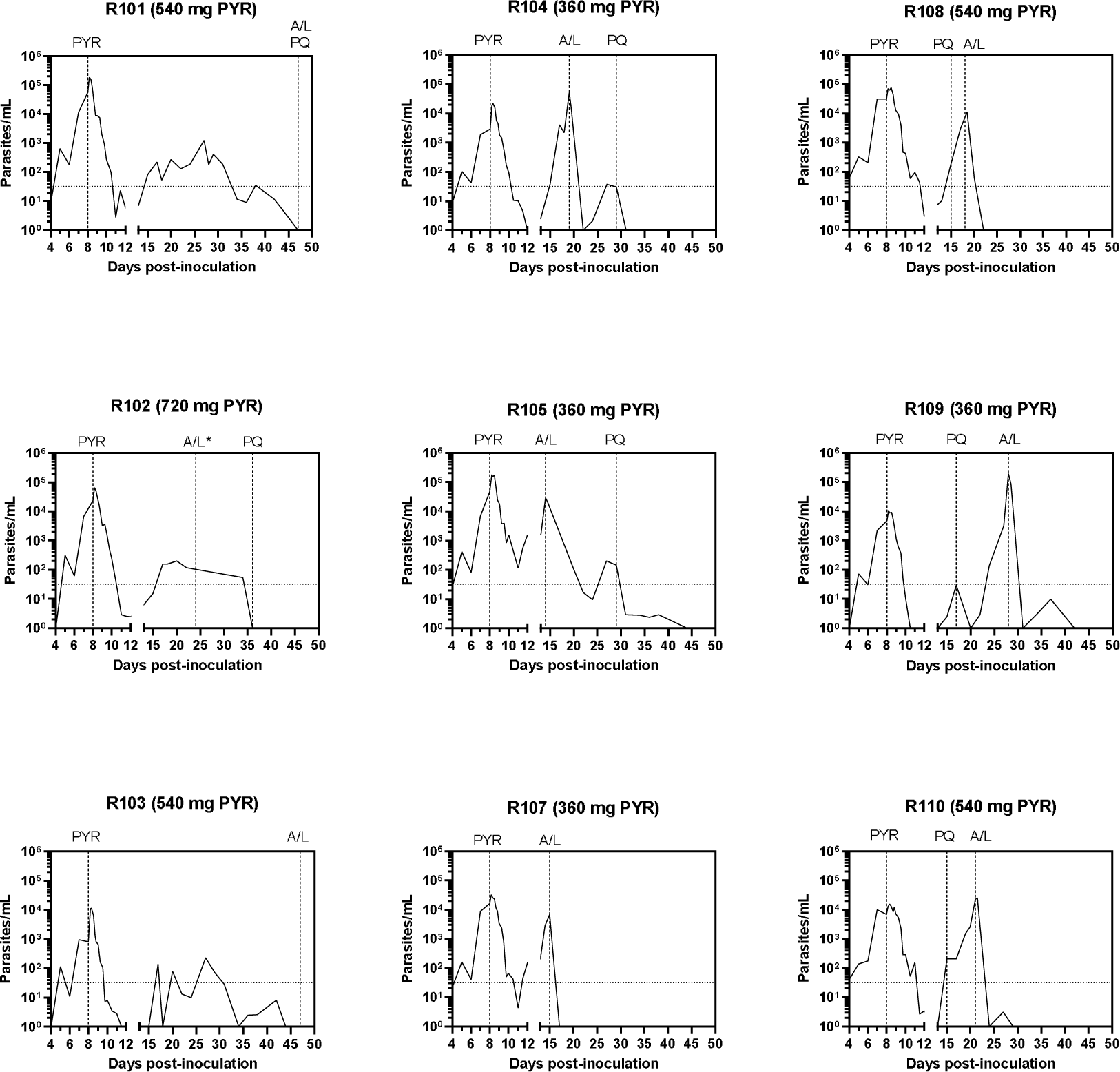
Individual participant parasitaemia-time profiles. Participants were inoculated intravenously with *P. falciparum*-infected erythrocytes on day 0 and were administered a single oral dose of 360 mg (n=4), 540 mg (n=4), or 720 mg (n=1) pyronaridine (PYR) on day 8. Definitive antimalarial treatment with a standard course of artemether-lumefantrine (A/L) was initiated in response to asexual parasite regrowth or on day 47. A single dose of primaquine (PQ) was administered if required to clear gametocytemia. Parasitaemia was measured using qPCR targeting the gene encoding *P. falciparum*18S rRNA. The horizontal dotted line indicates the lower limit of quantitation of the assay (32 parasites/mL) [27]. Data for participant R106 is not presented because the participant was not dosed with pyronaridine. *Definitive antimalarial treatment with artemether-lumefantrine was initiated on Day 24 for participant R102 because the participant tested positive for COVID-19 (asexual parasite regrowth was not observed at the time of treatment initiation).

**Figure 3.**
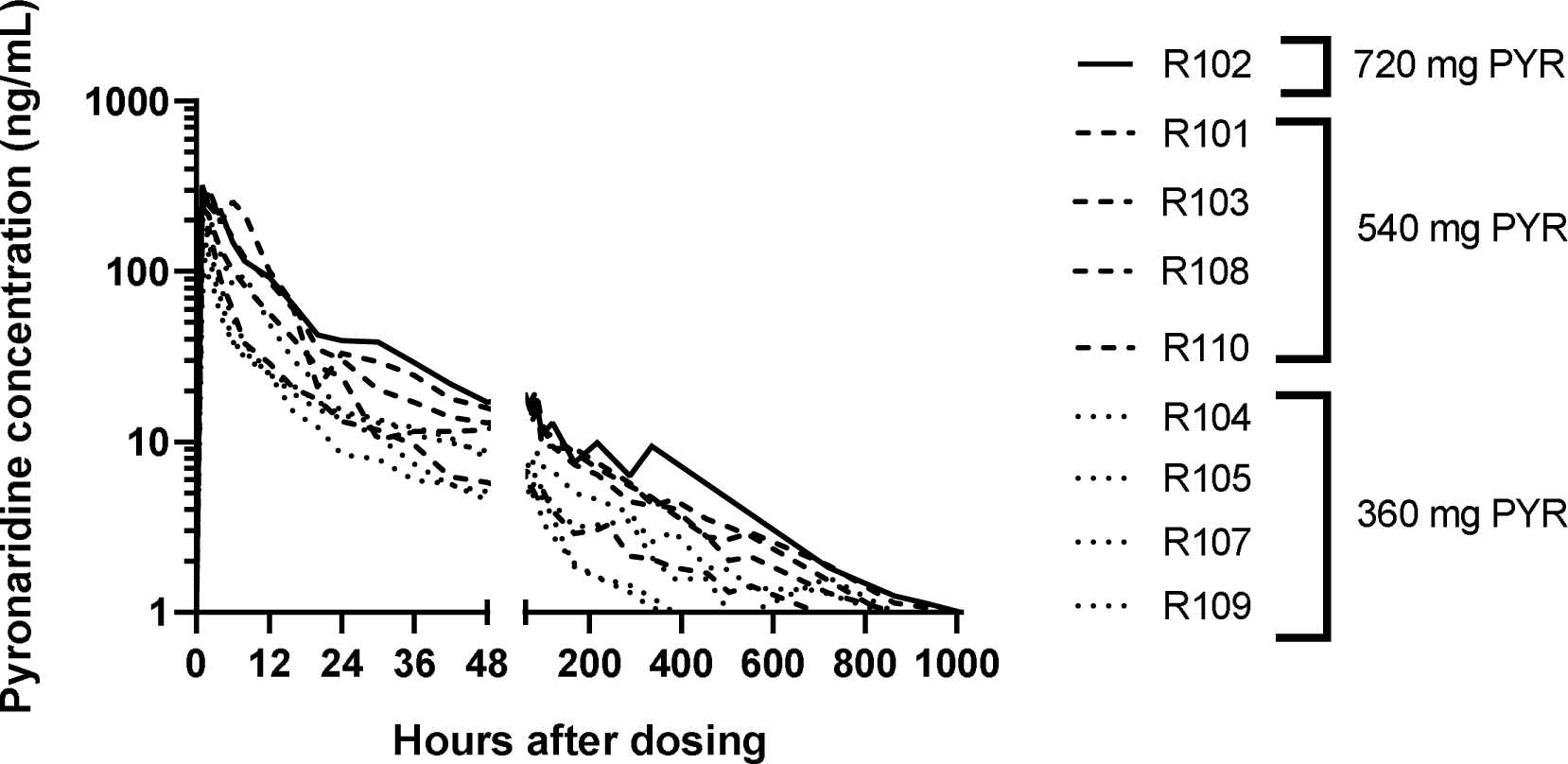
Individual participant whole blood pyronaridine concentration-time profiles. Participants were administered a single oral dose of 360 mg (n=4), 540 mg (n=4), or 720 mg (n=1) pyronaridine (PYR) 8 days following challenge with blood-stage *P. falciparum*. Whole blood pyronaridine concentration was measured using liquid chromatography-tandem mass spectrometry.

An initial reduction in parasitemia occurred in all participants following pyronaridine dosing, with a parasite clearance half-life of approximately 5 hours for each dose group (Table 2). Asexual parasite regrowth (differentiated from gametocytemia by qRT-PCR targeting parasite lifecycle specific transcripts; Figure S1) occurred in all 4 participants dosed with 360 mg (6-20 days post-dose) and in two participants dosed with 540 mg pyronaridine (10-13 days post-dose). Asexual parasite regrowth was not observed in the other two participants dosed with 540 mg up to day 48 when definitive antimalarial treatment with artemether-lumefantrine was initiated. The participant dosed with 720 mg pyronaridine tested positive for COVID-19 on day 24 (16 days post-dose) and thus received early definitive antimalarial treatment with artemether-lumefantrine on the same day. Asexual parasite regrowth was not observed in this participant at the time of treatment initiation.

**Table 2.** Parasite clearance parameters following single dose pyronaridine administration to healthy volunteers with induced *P. ciparum* parasitemia.

|  | Pyronaridine 360 mg<br>(N=4) | Pyronaridine 540 mg<br>(N=4) | Pyronaridine 720 mg<br>(N=1) |
| --- | --- | --- | --- |
| Log<sub>10</sub>PRR<sub>48</sub> (95% CI) | 2.58 (2.43 - 2.74) | 2.53 (2.37 - 2.68) | 2.85 (2.65 - 3.06) |
| PCt<sub>1/2</sub> [h (95% CI)] | 5.59 (5.28 - 5.94) | 5.71 (5.38 - 6.09) | 5.06 (4.72 - 5.46) |
| Parasite regrowth incidence [n (%)] | 4 (100) | 2 (50) | 0 (0) |

Log<sub>10</sub>PRR<sub>48</sub>: logarithm to base 10 of the parasite reduction ratio over a 48-hour period; PCt<sub>1/2</sub>: parasite clearance half-life.

**Table 3.** Pyronaridine venous whole blood non-compartmental pharmacokinetic parameters associated with single dose administration healthy volunteers with induced *P. falciparum* parasitemia.

|  | Pyronaridine 360 mg | Pyronaridine 540 mg | Pyronaridine 720 mg |
| --- | --- | --- | --- |
|  | (N=4) | (N=4) | (N=1) |
| C<sub>max</sub> (ng/mL) | 154.2 (43.0) | 250.6 (23.4) | 320.0 |
| t<sub>max</sub> (h) | 1.5 (1.0-2.0) | 2.0 (1.0-2.0) | 1.0 |
| AUC<sub>0-last</sub> (h×ng/mL) | 2452.5 (45.6) | 5202.4 (47.7) | 8776.1 |
| t<sub>1/2</sub> (h) | 233.0 (16.2) | 210.5 (11.0) | 189.8 |
| CL/F (L/h) | 123.6 (40.5) | 95.2 (41.7) | 79.3 |
| Vz/F (L) | 41533 (43.9) | 28921 (50.9) | 21711 |

Data for 360 mg and 540 mg groups are geometric means (coefficient of variation [%]) except t<sub>max</sub> which is median (range). Summary statistics were not calculated for the 720 mg group because N=1. C<sub>max</sub>: maximum observed concentration; t<sub>max</sub>: time to reach the maximum observed concentration; AUC<sub>0-last</sub>: area under the concentration-time curve from time 0 (dosing) to the last sampling time at which the concentration is at or above the lower limit of quantification; t<sub>1/2</sub>: apparent terminal half-life; CL/F: apparent total clearance; Vz/F: apparent volume of distribution.

Female and male gametocytes were detected in all participants after pyronaridine dosing (Figure S1). In cohort 1, a single dose of primaquine was administered to 4 participants after artemether-lumefantrine treatment to ensure the participants were free of gametocytes at the end of study (Figure 1). Given the relatively high level of gametocytemia observed in cohort 1, the study protocol was amended for cohort 2 to permit primaquine administration at any time after pyronaridine dosing. It was anticipated that earlier primaquine dosing would reduce the confounding effect of gametocytemia on the PK/PD analysis. Three of the four participants in cohort 2 were administered a single dose of primaquine prior to artemether-lumefantrine treatment (7-9 days after pyronaridine dosing). Rapid asexual parasite regrowth occurred in the fourth participant (7 days after pyronaridine dosing) obviating the need for primaquine dosing before artemether-lumefantrine treatment (Figure 1).

### Pharmacokinetic/Pharmacodynamic analysis

The PK of pyronaridine was described by a two-compartment model with zero-order absorption, linear elimination, and an additive and proportional residual error model (Table S1). Body weight at baseline was included in the structural model as a covariate using an allometric function with exponents fixed to 0.75 for clearance parameters and 1 for volume parameters. The individual observed pyronaridine concentrations were well described by the model (Figure S2). The estimated individual PK parameters were consecutively used in the PK/PD analysis of pyronaridine.

The relationship between pyronaridine blood concentrations and parasite killing was described by an E_max_ PK/PD model (Table S2). This model assumes an exponential growth of the parasites and a direct effect of concentrations on the rate of parasite killing/clearance. Population parameters were well estimated with a residual standard error (RSE) below 12%. RSE as well as shrinkage were below 30% for the estimated inter-individual variability. The individual observed parasitaemia profiles were well described by the model (Figure S3). Key efficacy parameters for pyronaridine derived from the final PK/PD model are presented in Table 4.

**Table 4.**
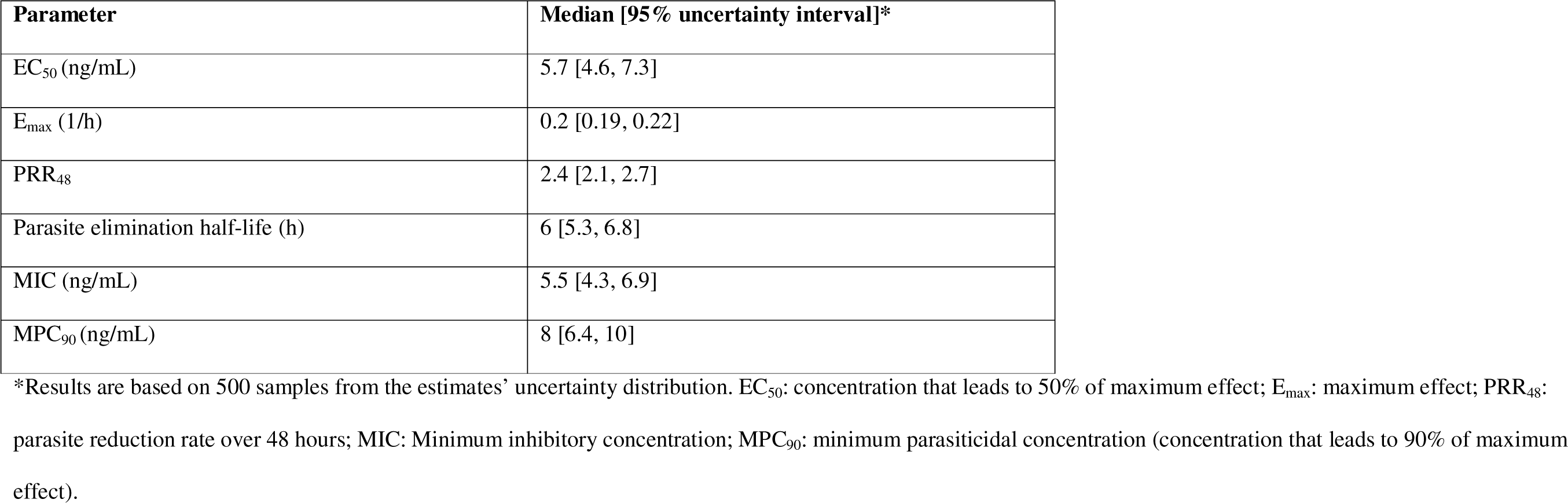
Key efficacy parameters for pyronaridine derived from the final PK/PD model.

### Safety and tolerability

A total of 139 AEs were reported for all 10 enrolled participants (Table 5), with 73 of these AEs being mild to moderate (grade 1 to 2) signs and symptoms of malaria (Table S4). There were no serious adverse events or adverse events with severity of grade 3 or higher. There were 14 AEs considered related to pyronaridine (Table S3), with gastrointestinal symptoms (abdominal pain, dyspepsia, gastroesophageal reflux, nausea, vomiting) the most frequent type of AE reported. Other AEs considered related to pyronaridine were headache, malaise, decreased appetite, and decreased hemoglobin. The event of decreased hemoglobin occurred in one male participant dosed with 360 mg pyronaridine, with a maximum decrease from baseline (pre-malaria challenge) of 30 g/L recorded 14 days after pyronaridine dosing. The participant’s hemoglobin level normalized spontaneously prior to the end of study visit. In addition to being considered potentially related to pyronaridine, the event was also considered potentially related to the malaria challenge and to blood sampling during the study. There were no clinically relevant trends in safety laboratory parameters over the course of the study, including liver function tests.

**Table 5.**
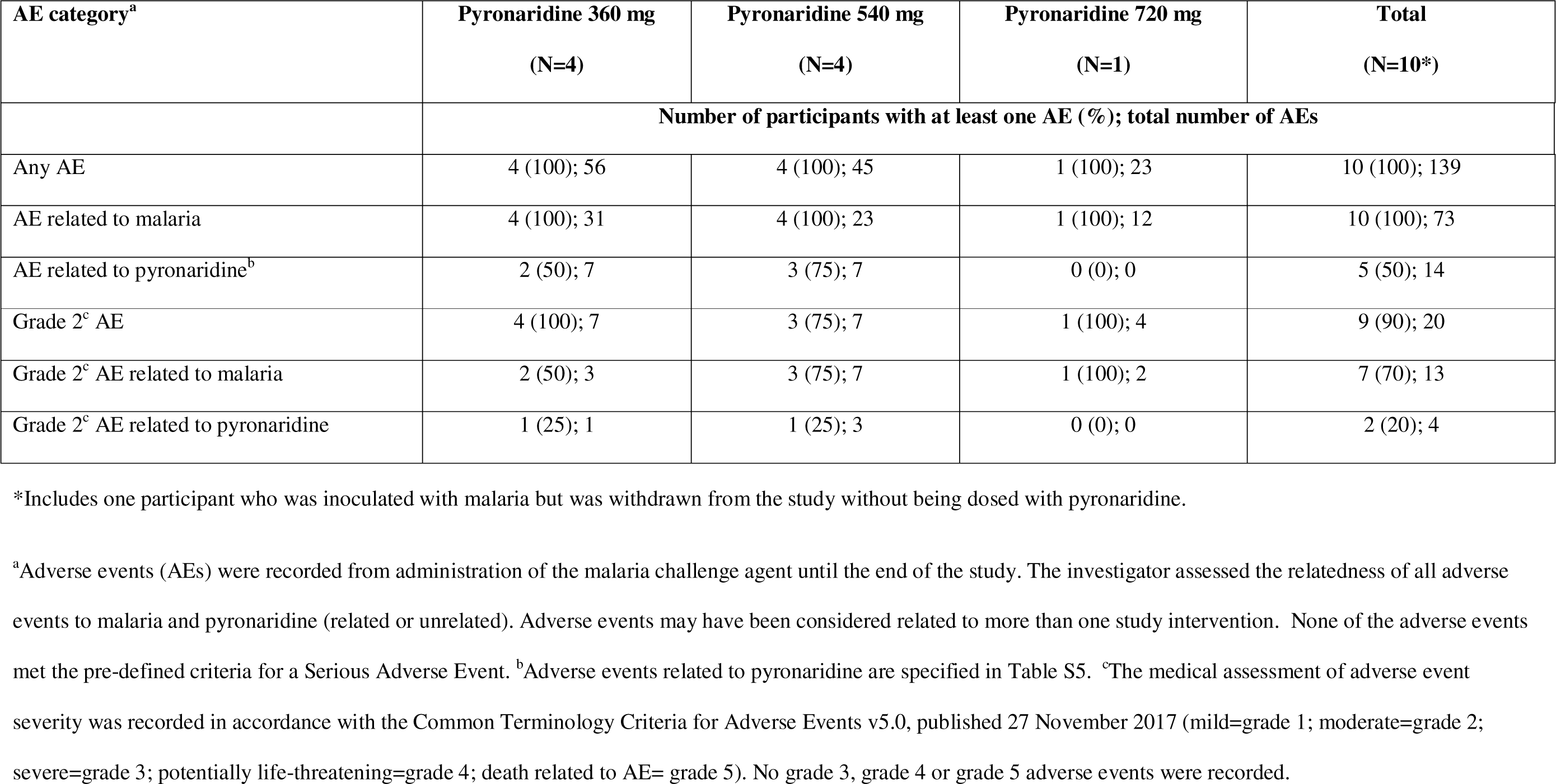
Summary of adverse events.

## DISCUSSION

The primary objective of this study was to characterize the antimalarial activity of pyronaridine by assessing the PK/PD relationship in malaria naïve adults using the *P. falciparum* IBSM model. Results confirmed that pyronaridine exhibits potent activity against blood-stage *P. falciparum*. The pyronaridine doses tested in this study (360 mg, 540 mg and 720 mg) all resulted in initial rapid parasite clearance, with a parasite clearance half-life of approximately 5 hours. There was no obvious difference in the rate of parasite clearance between dose levels, although this study was not powered to detect such differences. The parasite clearance rate characterized for pyronaridine is similar to that previously characterized for 10 mg/kg mefloquine (clearance half-life 6.2 hours) [12] and 640 mg piperaquine (clearance half-life 5.2 hours) [20] using the *P. falciparum* IBSM model. However, parasite clearance was slower than previously observed with 2 mg/kg oral artesunate (clearance half-life 3.2 hours) [21].

The PK profile of pyronaridine observed in this study is somewhat different to the PK of pyronaridine previously characterized in healthy adult volunteers and adult malaria patients administered the pyronaridine-artesunate combination. The elimination half-life was estimated to be 190-233 h (approximately 8-9 days) across the dose range in the current study as compared with 11.3 days (healthy volunteers) and 13.2 days (malaria patients) in individuals receiving combination therapy [5]. Differences in the populations used for analysis, and potential PK interactions between pyronaridine and artesunate, may be factors contributing to the different PK profiles observed. However, the small sample size of the current study in comparison with previous PK studies involving much larger sample sizes means it is not possible to draw definitive conclusions with respect to differences in PK profiles. Nevertheless, any potential differences in PK are not entirely relevant with respect to determining the PK/PD relationship of pyronaridine since the PK and PD measurements come from the same participants.

There were no safety concerns associated with single dose pyronaridine administration in this study, with mild to moderate gastrointestinal symptoms the most frequent adverse events observed. The good safety and tolerability profile of pyronaridine has been well established in several clinical trials involving both monotherapy and combination antimalarial therapy [5, 22, 23]. Additionally, the malaria-related adverse events observed were consistent with previous malaria volunteer infection studies.

A PK/PD model of pyronaridine was successfully developed in this study, enabling estimation of key efficacy parameters. It is anticipated that this model will be valuable in future efforts to expand the use of pyronaridine in new antimalarial combination therapies by informing partner drug selection and dosing considerations. At the time of writing, a phase 2a proof of concept study assessing the safety, efficacy, and PK of pyronaridine in combination with a new antimalarial candidate (M5717) in patients with acute uncomplicated *P. falciparum* malaria had begun recruiting (clinicaltrials.gov ID: NCT05689047). This study was initiated following a preclinical study of the combination *in vitro* and in the *P. falciparum* severe combined immunodeficient (SCID) mouse model [24]. The preclinical study indicated that the compounds had additive parasitological properties and that pyronaridine was able to suppress the selection of M5717-resistant parasites [24]. The fact that M5717 also appears to be active against transmissible gametocytes [25] likely adds to its suitability as a partner drug to pyronaridine given the high level of gametocytemia observed following administration of pyronaridine in the current study. We have previously conducted a VIS to characterize the antimalarial activity of M5717 using the IBSM model [26]. Thus, clinical PK/PD data is now available for both M5717 and pyronaridine, which will be valuable in the development of this antimalarial combination by informing dosing strategies. A combination VIS may also be worthy of consideration to characterize the PK/PD interaction of the drugs *in vivo*. Investigations are underway to identify other antimalarials that may represent good partner drugs to pyronaridine in a new combination therapy.

## Data Availability

All data produced in the present study are available upon reasonable request to the authors.

## Abbreviations

AEs: adverse events
BSM: induced blood stage malaria
PK/PD: pharmacokinetic/pharmacodynamics
VIS: volunteer infection study.

## ACKNOWLEWDGEMENTS

We thank all the volunteers who participated in the study, the University of the Sunshine Coast Clinical Trials Unit, Southern Star Research, Shin Poong Pharmaceutical for providing pyronaridine, the Queensland Paediatric Infectious Diseases laboratory for qPCR analysis, Swiss BioQuant for the measurement of pyronaridine concentrations, Jeremy Gower and Hayley Mitchell from the QIMR Berghofer Medical Research Institute for preparation of the malaria challenge inoculum, and Hanu Ramachandruni and Alice Neequaye from MMV.

## SUPPLEMENTARY MATERIAL

- Figure S1. Individual participant parasite lifecycle stage results.
- Table S1. Population parameter estimates of the final pharmacokinetic model of pyronaridine.
- Figure S2. Individual fit plots from the pharmacokinetic model of pyronaridine.
- Table S2. Population estimates of the final pharmacokinetic/pharmacodynamic model of pyronaridine.
- Figure S3. Individual fit plots from the pharmacokinetic/pharmacodynamic model of pyronaridine.
- Table S3. Incidence of adverse events related to pyronaridine.
- Table S4. Incidence of adverse events related to malaria.
- Text S1. Participant eligibility criteria.

## REFERENCES

1. World Health Organization. World Malaria Report 2022. Geneva.

2. Ashley EA, Dhorda M, Fairhurst RM, et al. Spread of artemisinin resistance in Plasmodium falciparum malaria. N Engl J Med 2014; 371(5): 411–23.

3. Balikagala B, Fukuda N, Ikeda M, et al. Evidence of Artemisinin-Resistant Malaria in Africa. N Engl J Med 2021; 385(13): 1163–71.

4. Hooft van Huijsduijnen R, Wells TN. The antimalarial pipeline. Curr Opin Pharmacol 2018; 42: 1–6.

5. Croft SL, Duparc S, Arbe-Barnes SJ, et al. Review of pyronaridine anti-malarial properties and product characteristics. Malaria journal 2012; 11: 270.

6. Bailly C. Pyronaridine: An update of its pharmacological activities and mechanisms of action. Biopolymers 2021; 112(4): e23398.

7. Agency EM. Pyramax: Opinion on medicine for use outside EU. Available at: https://www.ema.europa.eu/en/opinion-medicine-use-outside-EU/human/pyramax. Accessed 28 August 2023.

8. WHO Guidelines for malaria, 14 March 2023. Geneva: World Health Organization, 2023.

9. Burrows JN, Duparc S, Gutteridge WE, et al. New developments in anti-malarial target candidate and product profiles. Malaria journal 2017; 16(1): 26.

10. Barber BE, Abd-Rahman AN, Webster R, et al. Characterizing the blood stage antimalarial activity of tafenoquine in healthy volunteers experimentally infected with Plasmodium falciparum. Clin Infect Dis 2023.

11. Barber BE, Fernandez M, Patel HB, et al. Safety, pharmacokinetics, and antimalarial activity of the novel triaminopyrimidine ZY-19489: a first-in-human, randomised, placebo-controlled, double-blind, single ascending dose study, pilot food-effect study, and volunteer infection study. The Lancet Infectious diseases 2022; 22(6): 879–90.

12. McCarthy JS, Lotharius J, Ruckle T, et al. Safety, tolerability, pharmacokinetics, and activity of the novel long-acting antimalarial DSM265: a two-part first-in-human phase 1a/1b randomised study. The Lancet Infectious diseases 2017; 17(6): 626–35.

13. McCarthy JS, Ruckle T, Djeriou E, et al. A Phase II pilot trial to evaluate safety and efficacy of ferroquine against early Plasmodium falciparum in an induced blood-stage malaria infection study. Malaria journal 2016; 15: 469.

14. Phyo AP, Jittamala P, Nosten FH, et al. Antimalarial activity of artefenomel (OZ439), a novel synthetic antimalarial endoperoxide, in patients with Plasmodium falciparum and Plasmodium vivax malaria: an open-label phase 2 trial. The Lancet Infectious diseases 2016; 16(1): 61–9.

15. Llanos-Cuentas A, Casapia M, Chuquiyauri R, et al. Antimalarial activity of single-dose DSM265, a novel plasmodium dihydroorotate dehydrogenase inhibitor, in patients with uncomplicated Plasmodium falciparum or Plasmodium vivax malaria infection: a proof-of-concept, open-label, phase 2a study. The Lancet Infectious diseases 2018; 18(8): 874–83.

16. Rockett RJ, Tozer SJ, Peatey C, et al. A real-time, quantitative PCR method using hydrolysis probes for the monitoring of Plasmodium falciparum load in experimentally infected human volunteers. Malaria journal 2011; 10: 48.

17. Wang CYT, Ballard E, Llewellyn S, et al. Assays for quantification of male and female gametocytes in human blood by qRT-PCR in the absence of pure sex-specific gametocyte standards. Malaria journal 2020; 19(1): 218.

18. Tadesse FG, Lanke K, Nebie I, et al. Molecular Markers for Sensitive Detection of Plasmodium falciparum Asexual Stage Parasites and their Application in a Malaria Clinical Trial. Am J Trop Med Hyg 2017; 97(1): 188–98.

19. Marquart L, Baker M, O’Rourke P, McCarthy JS. Evaluating the pharmacodynamic effect of antimalarial drugs in clinical trials by quantitative PCR. Antimicrobial agents and chemotherapy 2015; 59(7): 4249–59.

20. Pasay CJ, Rockett R, Sekuloski S, et al. Piperaquine Monotherapy of Drug-Susceptible Plasmodium falciparum Infection Results in Rapid Clearance of Parasitemia but Is Followed by the Appearance of Gametocytemia. The Journal of infectious diseases 2016; 214(1): 105–13.

21. Watts RE, Odedra A, Marquart L, et al. Safety and parasite clearance of artemisinin-resistant Plasmodium falciparum infection: A pilot and a randomised volunteer infection study in Australia. PLoS Med 2020; 17(8): e1003203.

22. Tona Lutete G, Mombo-Ngoma G, Assi SB, et al. Pyronaridine-artesunate real-world safety, tolerability, and effectiveness in malaria patients in 5 African countries: A single-arm, open-label, cohort event monitoring study. PLoS Med 2021; 18(6): e1003669.

23. West African Network for Clinical Trials of Antimalarial D. Pyronaridine-artesunate or dihydroartemisinin-piperaquine versus current first-line therapies for repeated treatment of uncomplicated malaria: a randomised, multicentre, open-label, longitudinal, controlled, phase 3b/4 trial. Lancet (London, England) 2018; 391(10128): 1378–90.

24. Rottmann M, Jonat B, Gumpp C, et al. Preclinical Antimalarial Combination Study of M5717, a Plasmodium falciparum Elongation Factor 2 Inhibitor, and Pyronaridine, a Hemozoin Formation Inhibitor. Antimicrobial agents and chemotherapy 2020; 64(4).

25. Baragana B, Hallyburton I, Lee MC, et al. A novel multiple-stage antimalarial agent that inhibits protein synthesis. Nature 2015; 522(7556): 315–20.

26. McCarthy JS, Yalkinoglu O, Odedra A, et al. Safety, pharmacokinetics, and antimalarial activity of the novel plasmodium eukaryotic translation elongation factor 2 inhibitor M5717: a first-in-human, randomised, placebo-controlled, double-blind, single ascending dose study and volunteer infection study. The Lancet Infectious diseases 2021; 21(12): 1713–24.

27. Wang CYT, Ballard EL, Pava Z, et al. Analytical validation of a real-time hydrolysis probe PCR assay for quantifying Plasmodium falciparum parasites in experimentally infected human adults. Malaria journal 2021; 20(1): 181.

