## Supplementary Material for "Characterizing the blood stage antimalarial activity of pyronaridine in healthy volunteers experimentally infected with *Plasmodium falciparum*"

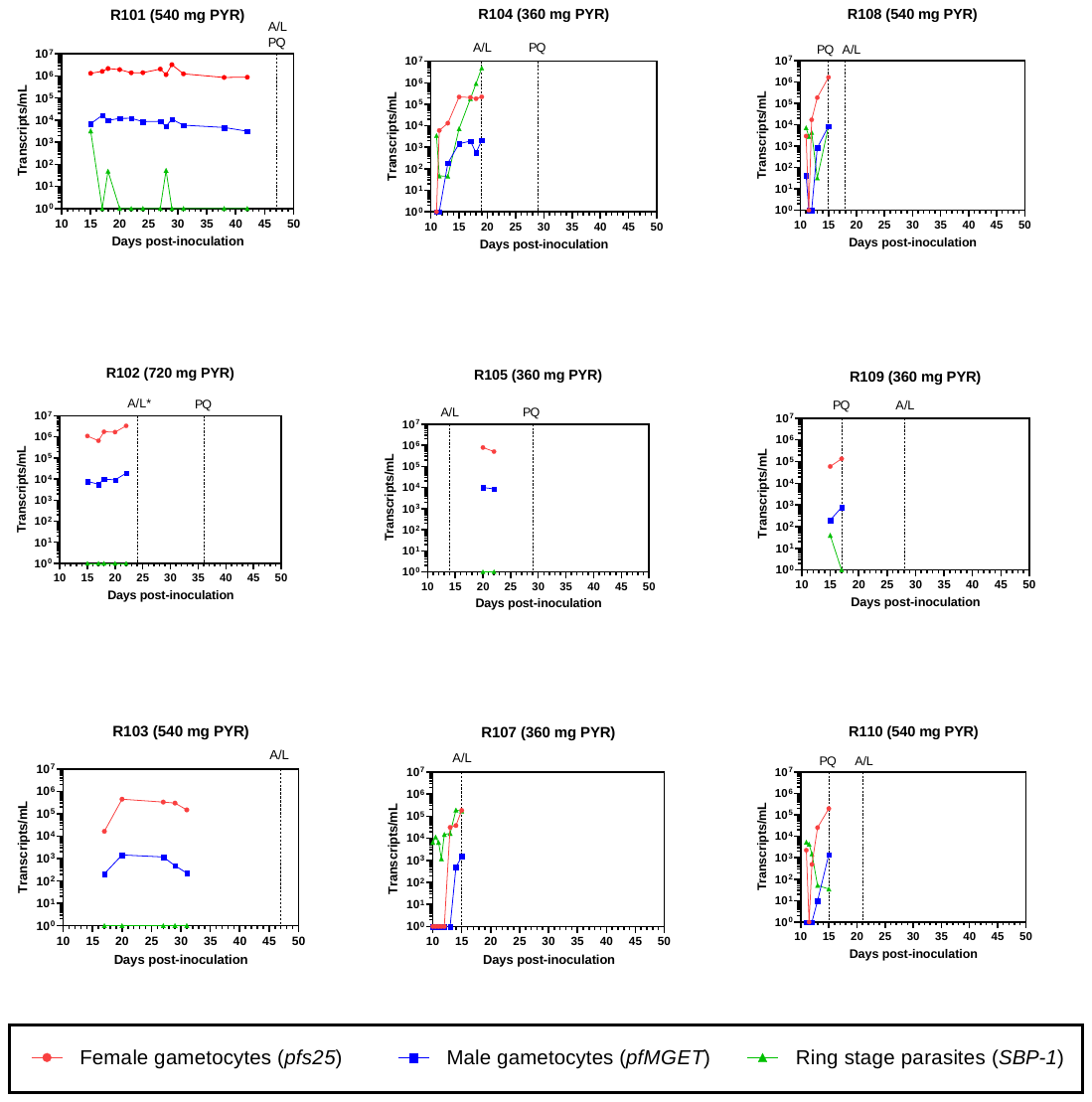

### **Figure S1. Individual participant parasite lifecycle stage results.**

The presence of female gametocytes, male gametocytes and ring stage parasites in participants’ blood samples was detected using qRT-PCR targeting *pfs25*, *pfMGET* and *SBP-1* parasite transcripts respectively. Definitive antimalarial treatment with a standard course of artemether-lumefantrine (A/L) was initiated in response to asexual parasite regrowth or on day 47. A single dose of primaquine (PQ) may have been administered after A/L treatment (cohort 1) or any time after pyronaridine dosing (cohort 2) to clear gametocytemia. *Definitive antimalarial treatment with artemether-lumefantrine was initiated on Day 24 for participant R102 because the participant tested positive for COVID-19 (asexual parasite regrowth was not observed at the time of treatment initiation).

### **Table S1. Population parameter estimates of the final pharmacokinetic model of pyronaridine**

| PARAMETER | VALUE | RSE | SHRINKAGE | COMMENT |
| --- | --- | --- | --- | --- |
| **Typical parameters** |  |  |  |  |
| Fabs0 | 1 (FIX) | - | - | Relative bioavailability (-) |
| CL | 48 | 8.34% | - | Apparent clearance (L/hour) |
| Vc | 897 | 9.22% | - | Apparent central volume (L) |
| Q1 | 81.5 | 10.9% | - | Apparent intercompartmental clearance (L/hour) |
| Vp1 | 6160 | 8.36% | - | Apparent peripheral volume (L) |
| Tk0 | 0.595 | 52.8% | - | Absorption time (hours) |
| Tlag1 | 0 (FIX) | - | - | Absorption lag time (hours) |
| **Inter-individual variability** |  |  |  |  |
| omega(Fabs0) | 0 (FIX) | - | - | LogNormal |
| omega(CL) | 0.365 | 66% | -5.2% | LogNormal |
| omega(Vc) | 0.254 | 27.1% | 1.6% | LogNormal |
| omega(Q1) | 0.305 | 26.2% | 2% | LogNormal |
| omega(Vp1) | 0.244 | 35% | 0.5% | LogNormal |
| omega(Tk0) | 0.738 | 52.9% | 33.4% | LogNormal |
| omega(Tlag1) | 0 (FIX) | - | - | Normal |
| **Parameter-Covariate relationships** |  |  |  |  |
| beta_CL(WT0) | 0.75 (FIX) | - | - | Body Weight in kg on CL (centered around: 55 kg) |
| beta_Vc(WT0) | 1 (FIX) | - | - | Body Weight in kg on Vc (centered around: 55 kg) |
| beta_Q1(WT0) | 0.75 (FIX) | - | - | Body Weight in kg on Q1 (centered around: 55 kg) |
| beta_Vp1(WT0) | 1 (FIX) | - | - | Body Weight in kg on Vp1 (centered around: 55 kg) |
| **Residual Variability** |  |  |  |  |
| error_ADD1 | 0.000223 | 22.8% | 4.74%* | Additive Error (ug/mL) - Compound concentration (ug/mL) |
| error_PROP1 | 0.171 | 6.88% | - | Proportional Error (fraction) - Compound concentration (ug/mL) |
| Objective function | -2003 | - | - | - |
| AIC | -1979 | - | - | - |
| BIC | -1977 | - | - | - |

Significant digits: 3 (Objective function rounded to closest integer value), omega values and error model parameters reported in standard deviation.
*Epsilon shrinkage (records with missing dependent variable and censored records not considered).

RSE: Residual Standard Error; AIC: Akaike information criterion; BIC: Bayesian Information Criterion.

### **Figure S2.** **Individual fit plots from the pharmacokinetic model of pyronaridine**

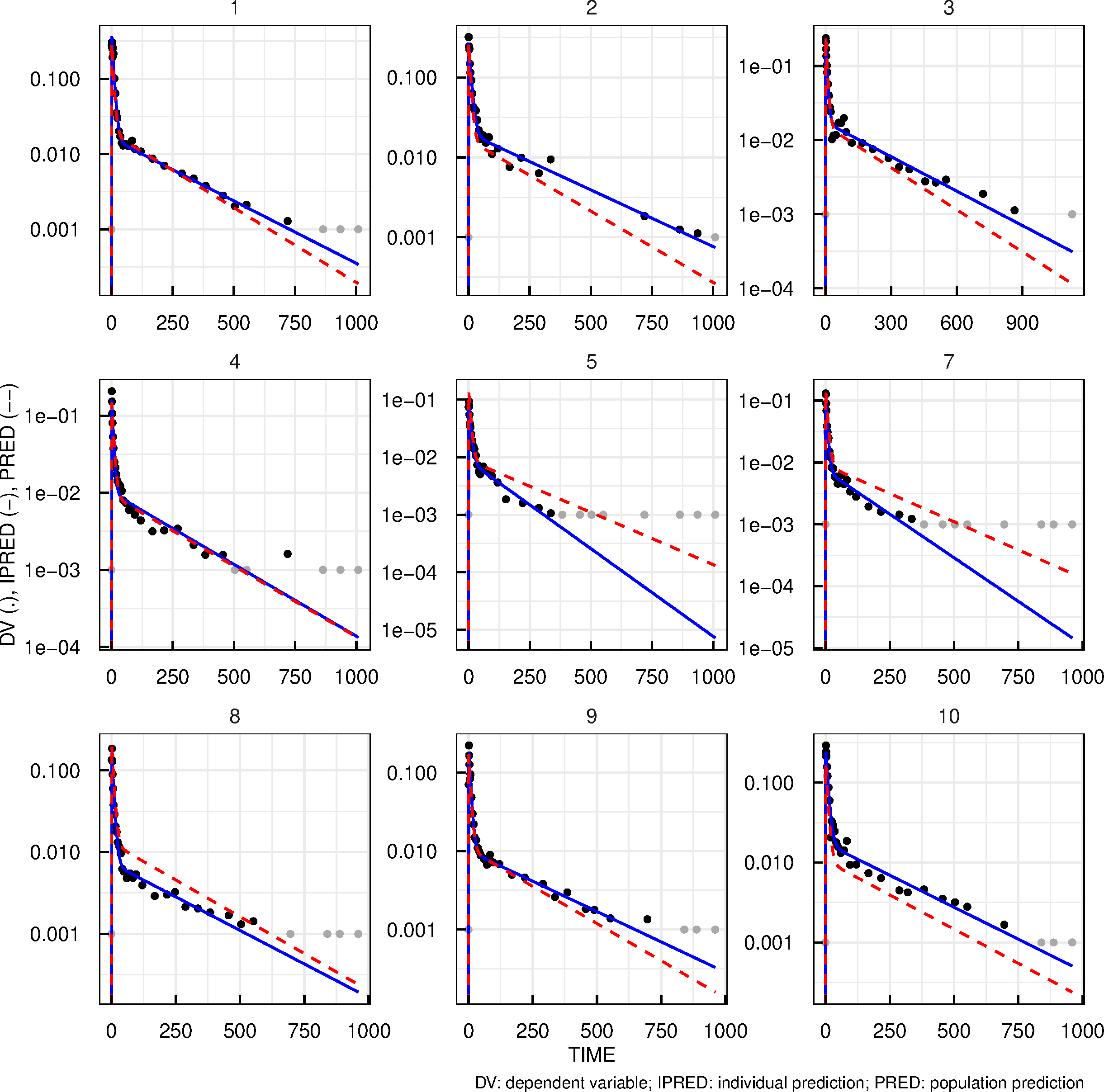

Blue continuous lines represent the individual model fit and red dashed lines represent the population model fit.

### **Table S2. Population estimates of the final pharmacokinetic/pharmacodynamic model of pyronaridine**

| PARAMETER | VALUE | RSE | SHRINKAGE | COMMENT |
| --- | --- | --- | --- | --- |
| **Typical parameters** |  |  |  |  |
| PLerr | -15 | 3.87% | - | Individual deviation from baseline parasitemia |
| GR | 0.0856 | 3.89% | - | Net parasite growth rate (1/hour) |
| EMAX | 0.201 | 3.87% | - | Maximum clearance rate (1/hour) |
| EC50 | 0.00575 | 11.3% | - | Concentration achieving 50percent of maximum effect (µg/mL) |
| hill | 6.62 | 2.42% | - | Hill coefficient (.) |
| **Inter-individual variability** |  |  |  |  |
| omega(PLerr) | 1.52 | 26.2% | 10.1% | Normal |
| omega(GR) | 0.1 (FIX) | - | 27.4% | LogNormal |
| omega(EMAX) | 0.0996 | 28.9% | 12.3% | LogNormal |
| omega(EC50) | 0.273 | 28.8% | 16.1% | LogNormal |
| omega(hill) | 0 (FIX) | - | - | LogNormal |
| **Residual Variability** |  |  |  |  |
| error_ADD1 | 1.02 | 3.36% | 9.9%* | Additive Error (log(p/ml)) - Log-transformed parasitemia (percent or 1/mL) |
| Objective function | 1626 | - | - | - |
| AIC | 1644 | - | - | - |
| BIC | 1646 | - | - | - |

Significant digits: 3 (Objective function rounded to closest integer value), omega values and error model parameters reported in standard deviation.
*Epsilon shrinkage (records with missing dependent variable and censored records not considered).

RSE: Residual Standard Error; AIC: Akaike information criterion; BIC: Bayesian Information Criterion.

### **Figure S3. Individual fit plots from the pharmacokinetic/pharmacodynamic model of pyronaridine**

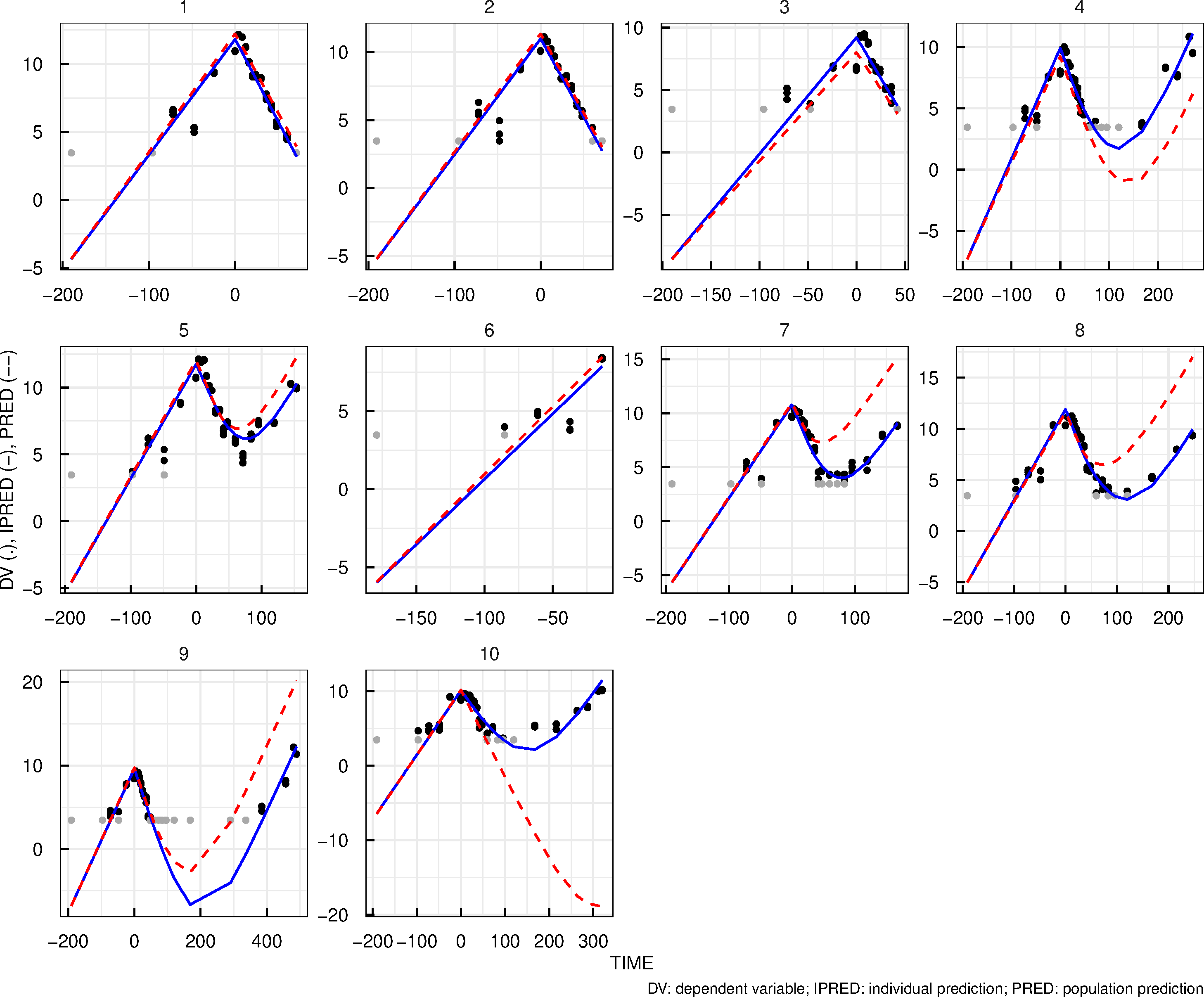

Blue continuous lines represent the individual model fit and red dashed lines represent the population model fit. Participant 6 (R106) was not dosed with pyronaridine.

### **Table S3.** **Incidence of adverse events related to pyronaridine**

| **Adverse event** | **Pyronaridine 360 mg  (N=4)** | **Pyronaridine 540 mg  (N=4)** | **Pyronaridine 720 mg  (N=1)** | **Total  (N=9^#^)** |
| --- | --- | --- | --- | --- |
|  | **Number of participants with at least one AE (%); total number of AEs** | | | |
| **Any AE related to pyronaridine*** | **2 (50); 7** | **3 (75); 7** | **0 (0); 0** | **5 (56); 14** |
| Abdominal pain | 1 (25); 1 | 1 (25); 1 | 0 (0); 0 | 2 (22); 2 |
| Dyspepsia | 0 (0); 0 | 1 (25); 1 | 0 (0); 0 | 1 (11); 1 |
| Gastroesophageal reflux disease | 0 (0); 0 | 1 (25); 1 | 0 (0); 0 | 1 (11); 1 |
| Nausea | 1 (25); 1 | 1 (25); 1 | 0 (0); 0 | 2 (22); 2 |
| Vomiting | 1 (25); 1 | 1 (25); 1 | 0 (0); 0 | 2 (22); 2 |
| Malaise | 1 (25); 1 | 0 (0); 0 | 0 (0); 0 | 1 (11); 1 |
| Haemoglobin decreased | 1 (25); 1 | 0 (0); 0 | 0 (0); 0 | 1 (11); 1 |
| Decreased appetite | 1 (25); 1 | 1 (25); 1 | 0 (0); 0 | 2 (22); 2 |
| Headache | 1 (25); 1 | 1 (25); 1 | 0 (0); 0 | 2 (22); 2 |

*All adverse events related to pyronaridine were either mild (grade 1) or moderate (grade 2) in severity as assessed using the Common Terminology Criteria for Adverse Events (CTCAE, version 5.0). ^#^Total excludes one participant who was not dosed with pyronaridine.

### **Table S4.** **Incidence of adverse events related to malaria**

| **Adverse event** | **Pyronaridine 360 mg  (N=4)** | **Pyronaridine 540 mg  (N=4)** | **Pyronaridine 720 mg  (N=1)** | **Total  (N=10^#^)** |
| --- | --- | --- | --- | --- |
|  | **Number of participants with at least one AE (%); total number of AEs** | | | |
| **Any AE related to malaria*** | **4 (100); 31** | **4 (100); 23** | **1 (100); 12** | **10 (100); 73** |
| Lymphopenia | 1 (25); 1 | 0 (0); 0 | 0 (0); 0 | 1 (10); 1 |
| Abdominal pain | 0 (0); 0 | 1 (25); 1 | 0 (0); 0 | 1 (10); 1 |
| Nausea | 0 (0); 0 | 1 (25); 1 | 1 (100); 2 | 2 (20); 3 |
| Vomiting | 0 (0); 0 | 1 (25); 1 | 0 (0); 0 | 1 (10); 1 |
| Chills | 1 (25); 3 | 1 (25); 1 | 1 (100); 1 | 3 (30); 5 |
| Fatigue | 2 (50); 3 | 2 (50); 3 | 1 (100); 2 | 6 (60); 9 |
| Feeling hot | 1 (25); 1 | 0 (0); 0 | 0 (0); 0 | 1 (10); 1 |
| Feeling of body temperature change | 1 (25); 1 | 0 (0); 0 | 0 (0); 0 | 1 (10); 1 |
| Malaise | 1 (25); 2 | 2 (50); 3 | 0 (0); 0 | 3 (30); 5 |
| Pyrexia | 1 (25); 4 | 3 (75); 4 | 1 (100); 2 | 6 (60); 11 |
| Haemoglobin decreased | 1 (25); 1 | 0 (0); 0 | 0 (0); 0 | 1 (10); 1 |
| Decreased appetite | 1 (25); 1 | 1 (25); 2 | 1 (100); 1 | 3 (30); 4 |
| Arthralgia | 0 (0); 0 | 1 (25); 2 | 0 (0); 0 | 2 (20); 3 |
| Back pain | 1 (25); 1 | 0 (0); 0 | 0 (0); 0 | 1 (10); 1 |
| Limb discomfort | 1 (25); 1 | 0 (0); 0 | 0 (0); 0 | 1 (10); 1 |
| Muscular weakness | 1 (25); 1 | 0 (0); 0 | 0 (0); 0 | 1 (10); 1 |
| Musculoskeletal chest pain | 1 (25); 1 | 0 (0); 0 | 0 (0); 0 | 1 (10); 1 |
| Myalgia | 2 (50); 3 | 1 (25); 1 | 0 (0); 0 | 4 (40); 5 |
| Dizziness | 1 (25); 1 | 0 (0); 0 | 0 (0); 0 | 1 (10); 1 |
| Headache | 3 (75); 6 | 2 (50); 3 | 1 (100); 4 | 7 (70);16 |
| Hyperhidrosis | 0 (0); 0 | 1 (25); 1 | 0 (0); 0 | 1 (10); 1 |

*All adverse events related to malaria were either mild (grade 1) or moderate (grade 2) in severity as assessed using the Common Terminology Criteria for Adverse Events (CTCAE, version 5.0). ^#^Total includes one participant who was not dosed with pyronaridine.

### **Text S1.** **Participant eligibility criteria**

**Inclusion Criteria**

Volunteers who met all of the following criteria were eligible for inclusion in the study:

**Demography**

1. Male or female (non-pregnant, non-lactating) aged 18 to 55 years inclusive who was contactable and available for the duration of the trial and up to two weeks following the EOS visit.
2. Total body weight greater than or equal to 50 kg, and a body mass index (BMI) within the range of 18 to 32 kg/m^2^ (inclusive). BMI is an estimate of body weight adjusted for height. It is calculated by dividing the weight in kilograms by the square of the height in metres.

**Health status**

3. Certified as healthy by a comprehensive clinical assessment (detailed medical history and full physical examination).
4. Fully vaccinated (meaning first, second and booster dose) against COVID-19 within 14 days of planned inoculation date.
5. Vital signs at screening (measured after 5 min in the supine position):
   - Systolic blood pressure (SBP) - 90–140 mmHg,
   - Diastolic blood pressure (DBP) - 40–90 mmHg,
   - Heart rate (HR) 40–100 bpm.
6. At Screening and pre-inoculation with the malaria challenge agent: normal standard mean of triplicate 12-lead electrocardiogram (ECG) parameters after 5 minutes resting in supine position in the following ranges:
   a. *QTcF ≤450 msec (male participants); QTcF ≤470 msec (female participants);*
   b. *QRS  50–120 msec*
   c. *PR interval ≤ 210 msec for both males and females, and*
   d. *Normal ECG tracing unless the PI or delegate considers an ECG tracing abnormality to be not clinically relevant.*
7. Women of childbearing potential (WOCBP) who anticipated being sexually active with a male during the trial must have agreed to the use of a highly effective method of birth control (see below) combined with a barrier contraceptive from the screening visit until 100 days after the last dose of pyronaridine (covering a full menstrual cycle of 30 days starting after 5 half-lives of last dose pyronaridine) and had a negative result on urine pregnancy test performed before inoculation with the malaria challenge agent.

*Note:*

a. *Highly effective birth control methods included: combined (oestrogen and progestogen containing) oral/intravaginal/transdermal/implantable hormonal contraception associated with inhibition of ovulation, progestogen-only oral/injectable/implantable hormonal contraception associated with inhibition of ovulation, intrauterine device, intrauterine hormone-releasing system, bilateral tubal occlusion, vasectomised partner, or sexual abstinence or same sex relationship.*
b. *Female participants who were abstinent (from penile-vaginal intercourse) must have agreed to start a double method if they started a sexual relationship with a male during the study. Female participants must have not be planning in vitro fertilisation within the required contraception period.*
8. Women of non-childbearing potential (WONCBP) were defined as:
a. *Natural (spontaneous) post-menopausal defined as being amenorrhoeic for at least 12 months without an alternative medical cause with a screening follicle stimulating hormone level (FSH) >25 IU/L (or at the local laboratory levels for post-menopause)*
b. *Premenopausal with irreversible surgical sterilization by hysterectomy and/or bilateral oophorectomy or salpingectomy at least 6 months before screening (as determined by participant medical history)*
9. Males who had, or may have had, female sexual partners of child bearing potential during the course of the study must have agreed to use a double method of contraception including condom plus diaphragm, or condom plus intrauterine device, or condom plus stable oral/transdermal/injectable/implantable hormonal contraceptive by the female partner, from the time of informed consent through to 70 days (covering a spermatogenesis cycle) after pyronaridine administration. Abstinent males must have agreed to start a double method if they began a sexual relationship with a female during the study and up to 70 days after the last dose of pyronaridine. Males with female partners of child-bearing potential that were surgically sterile, or males who had undergone sterilisation and had testing to confirm the success of the sterilisation, may also have been included and were not required to use above described methods of contraception.

**Regulations**

10. Completion of the written informed consent process prior to undertaking any trial-related procedure.
11. Must have been willing and able to communicate and participate in the whole trial.
12. Agreement to adhere to Lifestyle Considerations specified in the study protocol throughout trial duration
13. Agreement to provide current contact details (two telephone numbers including a mobile number and a responsible adult as an emergency contact) and a relevant email address.

**Exclusion Criteria**

Volunteers who met any of the following criteria were not eligible for inclusion in the study:

1. Individual who lived alone, OR, did not satisfy the following criteria: Participants who lived alone may have been included on a case-by-case basis, following discussion with the Principal Investigator.  Participants who lived alone must have identified and provided contact details of a support person who was aware of the participant’s participation in the study and was available to provide assistance if required (for example with contacting the participant in the event that study staff are unable to, or with transporting the participant to and from the study site if required).
2. Known hypersensitivity to pyronaridine, artesunate or any of its derivatives, artemether, lumefantrine or other artemisinin derivatives, proguanil/atovaquone, primaquine, or 4-aminoquinolines.
3. Haematology, biochemistry or urinalysis results at screening or at the eligibility visit that were outside of Sponsor-approved clinically acceptable laboratory ranges or were considered clinically significant by the PI or their delegate.
4. Participation in any investigational product trial within the 12 weeks preceding pyronaridine administration.
5. Symptomatic postural hypotension at screening (confirmed on two consecutive readings), irrespective of the decrease in blood pressure, or asymptomatic postural hypotension defined as a decrease in systolic blood pressure ≥20 mmHg within 2–3 min when changing from supine to standing position.
6. Any history of anaphylaxis or other severe allergic reactions including face, mouth, or throat swelling or any difficulty breathing, or other food or drug allergy that the Investigator considers may impact on participant safety. Participants with seasonal allergies/hay fever or allergy to animals or house dust mite that were untreated and asymptomatic at the time of dosing could be enrolled in the trial.
7. History of convulsion (including drug or vaccine-induced episodes). A medical history of a single febrile convulsion during childhood (< 5 years) was not an exclusion criterion.
8. Presence of current or suspected serious chronic diseases such as cardiac or autoimmune disease, diabetes, progressive neurological disease, severe malnutrition, hepatic or renal disease. Acute or progressive hepatic or renal disease, porphyria, psoriasis, rheumatoid arthritis, asthma (excluding childhood asthma, or mild asthma with preventative asthma medication required less than monthly), or epilepsy.
9. History of malignancy of any organ system (other than localised basal cell carcinoma of the skin or *in situ* cervical cancer), treated or untreated, within five years of screening, regardless of whether there is evidence of local recurrence or metastases.
10. Individuals with history of schizophrenia, bipolar disorder psychoses, disorders requiring lithium, attempted or planned suicide, or any other severe (disabling) chronic psychiatric diagnosis including generalised anxiety disorder.
11. Individuals who had been hospitalised within five years prior to enrolment for either a psychiatric illness or due to danger to self or others.
12. History of an episode of mild/moderate depression lasting more than 6 months that required pharmacological therapy and/or psychotherapy within the last 5 years; or any episode of major depression.

The Beck Depression Inventory-II (BDI-II) was used as a validated tool for the assessment of depression at screening. In addition to the conditions listed above, participants with a score of 20 or more on the BDI-II and/or a response of 1, 2 or 3 for item 9 of this inventory (related to suicidal ideation) were not eligible for participation. These participants were to be referred to a general practitioner or medical specialist as appropriate. Participants with a BDI-II score of 17 to 19 may have been enrolled at the discretion of an Investigator if they did not have a history of the psychiatric conditions mentioned in this criterion and their mental state was not considered to pose additional risk to the health of the participant or to the execution of the trial and interpretation of the data gathered.

13. History of recurrent headache (e.g. tension-type, cluster, or migraine) with a frequency of ≥2 episodes per month on average and severe enough to require medical therapy, during the 2 years preceding screening.
14. Presence of clinically significant infectious disease or fever (e.g., sublingual temperature ≥38.5°C) within the five days prior to inoculation.
15. Blood product donation to any blood bank during the 8 weeks (whole blood) or 4 weeks (plasma and platelets) prior to admission in the clinical unit on Day 8.
16. History or presence of alcohol abuse (alcohol consumption more than 40 g/4 units/4 standard drinks per day), or drug habituation, or any prior intravenous usage of an illicit substance.
17. Any individual who currently (within 14 days prior to inoculation) smokes >5 cigarettes/day.
18. Breastfeeding or lactating; positive serum pregnancy test at screening, positive urine pregnancy test upon admission or at other time points as specified by schedule of activities tables.
19. Any COVID-19 vaccine within 14 days of inoculation, any other vaccination within 28 days of IMP intake, and any vaccination planned up to the final follow-up visit.
20. Any corticosteroids, anti-inflammatory drugs (excluding commonly used over-the-counter anti-inflammatory drugs such as ibuprofen, acetylsalicylic acid, diclofenac), immunomodulators or anticoagulants within the past three months. Any individual currently receiving or having previously received immunosuppressive therapy (including systemic steroids, adrenocorticotrophic hormone or inhaled steroids) at a dose or duration potentially associated with hypothalamic-pituitary-adrenal axis suppression within the past year.
21. Use of prescription drugs (excluding contraceptives) or non-prescription drugs or herbal supplements (such as St John’s Wort), within 14 days or five half-lives (whichever was longer) prior to inoculation. Limited use of other non-prescription medications or dietary supplements, not believed to affect participant safety or the overall results of the trial, may have been permitted on a case-by-case basis following approval by the Sponsor in consultation with the PI. Participants were requested to refrain from taking non-approved concomitant medications from recruitment until the conclusion of the trial.
22. Cardiac/QT risk:

- Family history of sudden death or of congenital prolongation of the QTc interval or known congenital prolongation of the QTc interval or any clinical condition known to prolong the QTc interval.
- History of symptomatic cardiac arrhythmias or with clinically relevant bradycardia.

23. Any history of malaria or participation in a previous malaria challenge trial or malaria vaccine trial.
24. Must not have had malaria exposure that was considered by the PI or their delegate to be significant. This includes but was not limited to: history of having travelled to or lived (>2 weeks) in a malaria-endemic region during the past 12 months or planned travel to a malaria-endemic region during the course of the trial; history of having lived for >1 year in a malaria-endemic region in the past 10 years; history of having ever lived in a malaria-endemic region for more than 10 years inclusive.
25. Had evidence of increased cardiovascular disease risk (defined as >10%, 5-year risk for those greater than 35 years of age, as determined by the Australian Absolute Cardiovascular Disease Risk Calculator [http://www.cvdcheck.org.au/]). Risk factors include sex, age, systolic blood pressure (mm/Hg), smoking status, total and HDL cholesterol (mmol/L), and reported diabetes status.
26. History of splenectomy.
27. Individual unwilling to defer blood donations to the Blood Service for at least twelve months after the EOS visit.
28. Individual who has ever received a blood transfusion.
29. Any recent (<6 weeks) therapy with an antibiotic or drug with potential antimalarial activity (e.g. chloroquine, piperaquine phosphate, benzodiazepine, flunarizine, fluoxetine, tetracycline, azithromycin, clindamycin, doxycycline etc.).

**General conditions**

30. Any individual who, in the judgement of an Investigator, was likely to be non-compliant during the trial, or was unable to cooperate because of a language problem or poor mental development.
31. Any individual in the exclusion period of a previous trial according to applicable regulations.
32. Any individual who was an Investigator, research assistant, pharmacist, trial coordinator, or other staff thereof, directly involved in conducting the trial.
33. Any individual without good peripheral venous access.

**Biological status**

34. Positive result on any of the following tests: hepatitis B surface antigen (HBs Ag), anti-hepatitis B core antibodies (anti-HBc Ab), anti-hepatitis C virus (anti-HCV) antibodies, anti-human immunodeficiency virus 1 and 2 antibodies (anti-HIV1 and anti-HIV2 Ab).
35. Positive urine drug test. Any drug listed in the urine drug screen unless there was an explanation acceptable to an Investigator (e.g., the participant had stated in advance that they consumed a prescription or over-the-counter product that contained the detected drug) and the participant had a negative urine drug screen on retest by the pathology laboratory.
36. Severe G6PD deficiency.
37. Positive alcohol breath test.
38. Positive for SARS-CoV-2 by PCR or RAT.
39. Positive for red cell antibodies.
